# Prenatal and postpartum tobacco use in adolescents and young adults: a qualitative study

**DOI:** 10.64898/2026.09.10.26362664

**Authors:** Ajinkya Rai, Shinnyi Chou, Alison Sanfacon, Nicole Boss, Dahlia Lehman, Judy Chang, Natacha De Genna

## Abstract

**Background:** Younger obstetric patients are more likely to use combustible tobacco products and/or cannabis during pregnancy and, among those who quit, are more likely to relapse in the postpartum. Few studies have explored the context of prenatal tobacco use in this population from the narratives and perspectives of these patients. We performed a qualitative interview study of young pregnant people, their beliefs, attitudes, reasons for nicotine and tobacco use, and tobacco cessation experiences to address this gap.

**Methods:** Pregnant patients under the age of 22 were recruited from prenatal clinics in Pittsburgh for a cohort study. A subsample of participants were invited to participate in semi-structured interviews about tobacco and cannabis use. Two coders independently coded all transcripts using an open, inductive approach. The research team then used a qualitative description approach to perform thematic analysis by reviewing high frequency codes, identifying patterns and categories, and identifying themes.

**Results:** We conducted 57 interviews with 46 participants between the ages of 17-23 (M = 19.7 years). Most (n=33) identified as Black/African American, 9 as White, and 4 as Biracial. Thirty-eight were interviewed while pregnant (n=13 1^st^ trimester, n=18 2^nd^ trimester, n=12, 3^rd^ trimester) and 14 were interviewed in the postpartum. We identified the following themes: 1) participants recognized tobacco to be harmful for both the pregnant individual and the fetus; 2) they perceived tobacco use to be more harmful than cannabis use, and co-use to be more harmful than cannabis use alone; 3) peer and social influences contributed to tobacco initiation but also provided support for tobacco cessation; 4) they preferred to reduce or stop tobacco use through their own strategies rather than relying on clinicians or cessation aids; and 5) postpartum relapse was attributed to difficulty of dual transition into adulthood and parenthood.

**Conclusion:** While several themes – knowledge about the harms of tobacco use, greater perceived risk for tobacco compared to cannabis use, and the importance of peer and social support for tobacco cessation efforts – corroborate findings from studies with older pregnant patients, our study highlights some unique stressors contributing to postpartum relapse to combustible tobacco use in younger individuals and the need for better connection with clinical services supporting tobacco cessation.

## Introduction

Tobacco consumption during pregnancy is a significant public health concern, with an estimated 5.4% of pregnant individuals reporting cigarette use in 2021^1^. Prenatal tobacco exposure is linked to fetal growth restriction, placental abruption, preterm birth, low birthweight, orofacial clefts, and sudden infant death syndrome in addition to risks for childhood respiratory disease^2; 3^. However, most research on prenatal tobacco has focused on older populations, which may yield gaps in understanding the beliefs, attitudes, and experiences of younger individuals.

Despite declining prevalence overall, age-related disparities exist: young adults aged 20-24 years have the highest prevalence of prenatal smoking (10.7%), followed by adolescents aged 15-19 years^4^. Although 56% of individuals who smoke before pregnancy quit during fetal development, younger individuals (<20 years) are particularly susceptible to relapse postpartum^5; 6^. Concurrently, the sharpest rise in cannabis use during pregnancy is in populations of young individuals^7; 8^. Currently, evidence-based tobacco cessation interventions include behavioral counseling and financial incentives, which the US Preventive Services Task Force recommends offering to all pregnant individuals; evidence for nicotine replacement therapy is thus far inconclusive^9-11^. Universal interventions may not be as effective in younger pregnant patients, especially if they do not address cannabis use.

Younger pregnant individuals face distinct challenges; they transition into adulthood and parenthood simultaneously and feel strong influence from peers and social networks, particularly in relevance to substance use initiation and cessation^6; 12^. Qualitative methods are well-equipped to illuminate contextual factors shaping behaviors. To date, there has only been one qualitative study of tobacco use in pregnant adolescents. Constantine and colleagues focused on individuals who had successfully abstained for a month, many of their interviews were conducted postpartum, and the study was conducted at a time when there was less concurrent prenatal cannabis and e-cigarette use^13^. This study aims to prospectively explore beliefs, attitudes, and experiences related to tobacco use across pregnancy and in the postpartum among a more diverse sample of pregnant individuals aged 13-21 years through in-depth qualitative interviews.

## Methods

### Study Design

This qualitative study was part of a longitudinal, mixed-methods cohort study of prenatal tobacco and cannabis use in young pregnant individuals. The qualitative interviews aimed to examine participants’ experiences and perspectives about cannabis and tobacco use, especially their use across pregnancy and in the postpartum. Participants aged 13–21 with confirmed pregnancies were recruited from University of Pittsburgh Medical Center Magee-Womens Hospital antepartum unit, obstetric clinics, and affiliated neighborhood obstetrics-gynecology offices. Exclusion criteria included individuals not proficient in English and those currently using opioids or undergoing treatment for opioid use disorder. Once consented, all participants completed a baseline survey online, which collected details on demographics, substance use, and mental health. The University of Pittsburgh Institutional Review Board (IRB) approved all study procedures, and a separate IRB-approved consent was used for the qualitative interviews.

Participants in the parent study were eligible to participate in qualitative interviews if they had completed a baseline survey. They were contacted by research team members via phone, text, or email, and consent was reviewed with participants before interviews were conducted. We aimed to interview a sample of individuals during each trimester of pregnancy and 6 months or more postpartum. Initially, no prenatal tobacco or cannabis use was required to enroll in the qualitative study. Near the end of the parent study, we specifically recruited participants who had reported prenatal tobacco use in their baseline survey or tested positive for cotinine on a urine screen, to ensure that we had adequate representation of patients with a history of prenatal tobacco use in the qualitative study. Participants had the opportunity to complete up to four qualitative interviews: one in each trimester and one six to 18 months postpartum. Recruitment for the qualitative study continued until sampling was considered adequate, i.e., until enough individuals with tobacco use were interviewed at each time point and thematic saturation as determined through full-investigative-team review of the generated codes (see below).

One-on-one, semi-structured interviews were conducted between December 2020 and May 2024 and audio-recorded through a HIPAA-compliant version of Zoom, or by telephone with a digital recorder. Interviews lasted approximately 45–60 minutes and recordings were labeled with a participant ID number. Interview guides were modeled after a prior qualitative study of the beliefs and attitudes of adult pregnant women who used cannabis^14^. Trimester-specific guides included questions relevant to that time period of pregnancy, cessation strategies and experiences, information sources, interactions with obstetric care providers, and recommendations from participants for future substance-use research and intervention (Table 1). Interviewers (a medical student, research project specialists, and a research coordinator) received intensive training in qualitative interviewing, including several practice interviews from an obstetrician-gynecologist with extensive expertise in conducting and teaching qualitative research. Interviewees were paid $20 following completion of each interview.

**Table 1.** Interview Guide Topics.

| **Topic** | **Sample Questions** |
| --- | --- |
| Pregnancy experience | • How is your pregnancy going so far?<br>• How are you feeling?<br>• Is your pregnancy going as you expected? |
| Substance use | • Has your pattern of use changed since you found out you were pregnant (or since your last interview)?<br>• How old were you when you first tried cannabis/tobacco? Tell me about that experience.<br>• Tell me about why you are currently using cannabis/tobacco.<br>• Do you think your tobacco use is related to your cannabis use? If so, how are they related? If no, what separates them?<br>• Are you trying to change your cannabis or tobacco use currently? If so, how, and what has that process been like?<br>• Have you found other coping mechanisms besides cannabis/tobacco?<br>• Since we last spoke, have there been any incidents or events that led you to use? |
| Beliefs and attitudes | • What do you think about the use of cannabis/tobacco during pregnancy?<br>• Do you think one is safer than the other?<br>• What are the good/bad things related to cannabis and tobacco use during pregnancy?<br>• What are the effects of using cannabis/tobacco during pregnancy?<br>• Where do young, pregnant people get information about cannabis and tobacco?<br>• What do you think would help young, pregnant people get more information on these topics?<br>• After your baby is born, what are your plans for cannabis and tobacco use? |
| Legal/CYF involvement, perceptions | • What have your health care providers or social workers told you about what will happen if your baby tests positive for cannabis at birth?<br>• What impact do legal issues or CYF involvement have on your decision to use cannabis close to the time you are going to deliver?<br>• How has the legalization of medical cannabis impacted your views on cannabis use? |
| Obstetric care experience | • Did your doctor or another provider talk to you about cannabis and/or tobacco use during your appointment?<br>• How do you feel about that interaction?<br>• What has influenced your decision to talk or not talk to your doctor about cannabis or tobacco use?<br>• What can healthcare providers do to help young, pregnant people feel more comfortable talking to them about cannabis and tobacco use?<br>• At your appointment, were you provided with information about the use of cannabis or tobacco during pregnancy? |

### Reflexivity

The authors recognize that aspects of their identities, personal experiences, relationships with each other, and interactions with participants can influence perceptions and choices throughout the design, data collection, and analysis processes. The interview and coding team consisted of all cisgender females with interest in women’s mental health. All team members received dedicated training on the conduct of qualitative interviews with a well-known expert in the field and completed mock interview sessions prior to interviewing participants. The analysis and writing team consisted of the same individuals with the addition of one cisgender male medical student pursuing psychiatry. There were no previously established relationships between the participants and team members other than through participation in the prior quantitative study sessions of the larger parent study.

### Data Analysis

Verbatim transcription of the first 13 interviews were completed by study personnel, through manual editing of the automated Zoom-or Word-generated transcripts to match the audio recordings. Landmark Associates, Inc., was contracted to complete transcriptions of the remaining interviews, and all transcripts were reviewed by interviewers for accuracy. We used a qualitative descriptive approach to perform thematic analysis as we sought to adhere closely to the words and narratives of our participants without any predetermined framework or theory^15-17^.

To create the qualitative codebook, two researchers independently applied an open, iterative coding process to transcripts using the NVivo software (version 14) to store and organize the codes. Iterations involved first immersing in a few transcripts by reading them through in their entirety to familiarize and observe recurring patterns without coding, followed by reviewing the same transcripts again and generating novel codes reflective of what participants shared. These codes were then compared to each additional transcript content to determine whether codes should be retained, merged, split, or eliminated. The coders met regularly to review coding decisions using a constant-comparison method (Crabtree & Miller, 1999), and disagreements were explored in detail to determine consensus. Once transcripts stopped producing novel codes (i.e., an indication that saturation had been reached), the finalized codebook underwent team review before being applied to all transcripts.

Subsequent coding was completed by the same two researchers, first independently coding individual transcripts, then meeting to review and compare decisions to ensure consensus. All coded interviews were reviewed at regular full-investigative-team meetings, where the coding process and definitions were reviewed and emerging themes considered. After all transcripts were coded, coders reviewed high frequency codes, discussed patterns and categories, and together with the full investigative team, identified themes and subthemes.

## Results

### Subject Characteristics

Forty-six participants consented to the qualitative portion of the study. Thirty-eight completed at least one prenatal interview (n= 13, 1^st^ trimester, n = 18, 2^nd^ trimester, n = 12, 3^rd^ trimester), and 14 completed a postpartum interview. Six participants completed two interviews across different pregnancy and postpartum time points. Participants ranged in age from 17 to 23 years, with the largest percentage (n=15, 33%) being 21 years old (Table 2). Most identified as Black or African American (n=33, 72%) and lived with their parents at the time they completed their baseline surveys (n=19, 41%). Eight (17%) participants were students. Fourteen (30%) worked either part-or full-time. Most participants were expecting their first child (n=30, 65%).

**Table 2.** Characteristics of Study Participants.

| **Characteristic** |  | **N (%)** |
| --- | --- | --- |
| Age |  |  |
|  | 17 | 3 (6%) |
|  | 18 | 9 (20%) |
|  | 19 | 7 (15%) |
|  | 20 | 11 (24%) |
|  | 21 | 15 (33%) |
|  | 23 | 1 (2%) |
| Race |  |  |
|  | Black | 33 (72%) |
|  | White | 9 (20%) |
|  | Biracial | 4 (8%) |
| Self-Reported Tobacco Use in Baseline Survey |  |  |
|  | Never used | 22 (48%) |
|  | Stopped when learned of pregnancy | 11 (24%) |
|  | Once a month | 1 (2%) |
|  | Once a week | 0 (0%) |
|  | 2–3 days a week | 3 (6%) |
|  | 4 or more days a week | 3 (6%) |
|  | Daily | 6 (13%) |
| Self-Reported Cannabis Use in Baseline Survey |  |  |
|  | Never used | 13 (28%) |
|  | Stopped when learned of pregnancy | 18 (39%) |
|  | One a month | 2 (4%) |
|  | Couple times a month | 1 (2%) |
|  | Once a week | 3 (6%) |
|  | 2–3 days a week | 4 (8%) |
|  | 4 or more days a week | 1 (2%) |
|  | Daily | 4 (8%) |
| Reproductive history |  |  |
|  | First child | 30 (65%) |
| Education and employment |  |  |
|  | Enrolled in school | 8 (17%) |
|  | Employed full time | 7 (15%) |
|  | Employed part time | 7 (15%) |
|  | Unemployed | 18 (39%) |
| Living arrangement |  |  |
|  | Living with parents | 19 (41%) |
|  | Living alone (with child/ren) | 1 (2%) |
|  | Living with father of baby | 12 (26%) |
|  | Living with another relative | 3 (8%) |
|  | Other living arrangement | 2 (5%) |

In the baseline survey assessing frequency of cannabis and tobacco use (Table 2), many participants reportedly stopped their tobacco use when they found out they were pregnant (n = 11, 24%). However, more participants (n = 12, 26%) reported ongoing tobacco use at a frequency of at least once a week. Even more (n = 15, 33%) reported prenatal cannabis use. There were no significant differences in baseline characteristics (age, race, employment, school enrollment, gravidity, or prenatal cannabis use status) between individuals who participated in a qualitative interview versus those who completed a baseline survey but were not enrolled in this portion of the study.

### Themes

We identified five major themes related to prenatal and postpartum tobacco use in our analysis from the participants: 1) participants recognized tobacco to be harmful for both the pregnant individual and the fetus; 2) they perceived tobacco use to be more harmful than cannabis use, and co-use to be more harmful than cannabis use alone; 3) peer and social influences contributed to tobacco initiation but also provided support for tobacco cessation; 4) they preferred to reduce or stop tobacco use through their own strategies rather than relying on clinicians or cessation aids; and 5) postpartum relapse was attributed to difficulty of dual transition into adulthood and parenthood.

#### Tobacco is harmful for both the pregnant individual and the fetus

Participants expressed strong beliefs that tobacco is harmful for both the pregnant individual and the pregnancy/offspring (negative consequences of tobacco use appeared in 12 interviews; negative beliefs regarding tobacco use reported in 37 interviews; tobacco risks in pregnancy expressed in 39 interviews). For individuals who had no lifetime history of tobacco use, they expressed concerns regarding bad odor, addiction, general health effects (e.g., on skin, oral health, hair quality), cancer and death as reasons for never using: “I really never heard anything good about it, they always say don’t ever do it because it gets you addicted. People have been dying from it– like the vapes or stuff like that, so I guess, I really never heard anything good about it. It’s all negative” (Age range 21-25, never used tobacco).

Similarly, for individuals who stopped tobacco use upon learning about their pregnancy, they described having always identified tobacco as harmful even prior to pregnancy and prior to discontinuation (e.g., coughing, asthma, chest pain, stomach pain, headaches, nausea, bad taste), with the additional concern that tobacco use during pregnancy would be harmful to not only themselves, but also their pregnancy and offspring: “It’s not good, it’s not good, it’s not good at all because, like… I mean when I smoke cigarettes after I have our first kid it took me, maybe like two months to start smoking cigarettes again, but I never would smoke them whole because they’re like nasty so like I was in this pregnancy, when I found that I was pregnant I quit smoking cigarettes, maybe like two, three days later it’s like it just happened, I don’t think cigarettes are good” (Age range 21-25, quit tobacco at pregnancy recognition).

Even participants who continued to use tobacco during pregnancy described beliefs that tobacco may be harmful during pregnancy: “I think maybe it could have an impact on whether the baby has asthma, because I am smoking with the baby growing inside of me. If any of that nasty cigarette smoke gets somehow down near the baby, which I don’t see how it would even get down there—but if somehow it did, it could affect the baby’s lungs and give them asthma or a lung condition. I don’t want that to happen. That’s why, when I go for my ultrasounds, I make sure that they check everything just so I know that I’m not hurting my baby” (Age range 16-20, tobacco use during pregnancy).

In line with the belief that tobacco is harmful for both the pregnant individual and the fetus, many participants expressed the belief that people should not use tobacco during pregnancy: “Okay, I think that’s a big no because, um, it can affect your pregnancy. It can affect your baby and I don’t, I don’t think that’s a good idea at all. I feel like that’s a big no. Once you find out like once you’re confirmed pregnant, your pregnancy is confirmed, I feel like the tobacco and alcohol should be a big no” (Age range 16-20, quit tobacco at pregnancy recognition). Whether they continued to use tobacco during pregnancy or not, many participants expressed the belief that individuals who used tobacco during pregnancy were likely experiencing significant difficulties with cessation despite a desire to quit or cut down: “I don’t necessarily think vaping [nicotine] is good for pregnancy, I will say depending on how much nicotine you use. Cigarettes should be a no, but there’s some people who do have a hard time getting off of them” (Age range 16-20, tobacco use during pregnancy). Some participants who used tobacco during pregnancy additionally described experiencing adverse reactions to tobacco during this time period: “like my first pregnancy, I stopped smoking at 14 weeks, but I found out, I was pregnant at… maybe… 11. So I stopped smoking at 14 weeks, and it will make me nauseous or give me bad headaches. Even when somebody smoked it around me like I would throw up” (Age range 21-25, quit tobacco at pregnancy recognition).

Interestingly, participants described most of these beliefs as things they learned passively through conversations, rather than information they specifically sought out related to pregnancy. The negative consequences and harm related to general health appear to be common knowledge, with some participants even discussing terms such as “carcinogen” or referencing second-hand smoke and the delayed adverse effects later in life. Adverse effects toward infants were less specifically described, though some individuals did specifically raise concerns regarding fetal development and childbirth complications.

#### Tobacco use is more harmful than cannabis use, and co-use is more harmful than cannabis use alone

Participants explicitly described tobacco as worse than cannabis. Many participants shared the beliefs that tobacco is more harmful, harsher, more addictive, and more physically damaging than cannabis. In contrast, cannabis was viewed as less problematic in general, especially when regarding the addictive nature of the two substances: “Thinking about quitting tobacco was harder than thinking about quitting marijuana. I feel like it’s more addictive to smoke tobacco than it is to smoke marijuana for me personally” (Age range 21-25, tobacco use during pregnancy).

Similarly, when contrasting the use of the two substances during pregnancy, participants described the belief that tobacco exposure is more harmful, while cannabis may be acceptable: “I don’t think people should smoke cigarettes while they are pregnant just because tobacco is more harsher and stronger than marijuana. And it’s-it’s-it’s already bad for you, regardless, bad for your lungs, bad for your body period. So, while being pregnant, if it’s bad for your lungs then it’s bad for the baby’s lungs. Like something like that. Marijuana, unless it’s laced, there’s nothing wrong with it. Like you can’t-you can’t get sick from it. It can’t mess up your body without nothing being in it” (Age range 16-20, never used tobacco). This belief was echoed by both individuals with and without tobacco use history: “Before I got pregnant, I was smoking ‘em [tobacco] a lot, like heavy. After the pregnancy, I really don’t care. That’s ‘cause I be smoking weed, for real, so I don’t care. That’s ‘cause, back then, I didn’t have weed. Basically, cigarettes was around more than weed back before I got pregnant, but now, as I get older and I got my own money and stuff like that, weed is more around than cigarettes is, so I really don’t care for cigarettes and stuff. I feel like that’s a good thing ‘cause me, to be honest, I feel like cigarettes is way worse than smoking weed” (Age range 16-20, quit tobacco at pregnancy recognition).

In addition, some participants expressed the belief that tobacco and cannabis co-use is more harmful than cannabis use alone. They attribute this to the negative effects of tobacco, while describing cannabis use alone as less likely to cause harm during pregnancy.

#### Peer and social influences contributed to tobacco initiation but also provided support for tobacco cessation

Participants with a history of tobacco use often stated the reason for use as related to peer and social situations. This was often mentioned when describing initiation of tobacco use, while later, ongoing use was described as a method of coping with stress and boredom or related to addiction. Some participants also described family influences as the primary factor that led to tobacco use initiation, noting that growing up observing family members using tobacco normalized the activity. Importantly, participants described that although adult family members may voice initial disapproval, they sometimes would become accepting of the participants’ tobacco use, even sharing tobacco: “It started with me sneaking cigarettes from my mom when she was asleep, and I pretty much have just been smoking cigarettes since then… she was very mad at first, and then she started buying cigarettes for me… Anytime I see her, we openly smoke” (Age range 16-20, tobacco use during pregnancy).

However, many participants also reported family members or partners as a main influence in their decision making. These individuals urged them to quit tobacco use during pregnancy, citing concerns about fetal health, and sometimes went as far as directly refusing to provide tobacco or financial means to obtain tobacco. Participants described their social circles as generally supportive of their efforts to quit or cut down. This includes mentions of support from grandparents, parents, partners and peers: “Throughout the last pregnancy with my son and this current pregnancy, I have found that I have cut down a lot. I have learned to talk it out with a partner, my partner, and/or a family member that can help me” (Age range 21-25, quit tobacco at pregnancy recognition). Sometime this also came in the form of friends or family members avoiding tobacco use in the presence of the participants, or providing ongoing emotional support and encouragement as participants reduced or abstained from tobacco use.

In contrast, participants perceived healthcare providers as less helpful. They described provider communications regarding tobacco use and cessation as brief, sometimes offering cessation aids without offering in-depth discussions about strategies for success: “I wouldn’t say it was really talked about… I told ‘em, like, I don’t use, uh, marijuana, and I smoked tobacco, like, here and there, but not really. They just, kinda just went on to the next subject of the appointment” (Age range 16-20, quit tobacco at pregnancy recognition). Another participant shared: “they tried to give me the patches and they tried to prescribe the-told me to do the nicotine gum, and all this other stuff and nothing was working” (Age range 21-25, tobacco use during pregnancy).

#### Reducing / stopping tobacco use through individual strategies rather than relying on clinicians or cessation aids

The majority of participants who identified as using tobacco prior to pregnancy reported reducing or quitting tobacco use upon pregnancy recognition, and no one maintained the same level of tobacco use once they decided to continue with the pregnancy course. Most participants described health concerns as the primary motivation, both related to intolerance of tobacco during pregnancy (e.g., increased headache and nausea associated with tobacco use) and fear of harm to the developing offspring. These individuals devised their own strategies for reducing or stopping use: “I do smoke like half a cigarette in the morning, and then I’ll wait ‘til night and finish that cigarette. That’s just my process to slowly quitting because it was worse” (Age range 16-20, tobacco use during pregnancy). Some participants expressed that keeping themselves occupied with other activities helped with intentionally extending the time between tobacco use throughout the day and week “before I quit for about three days, I was taking a couple hits, putting it out, take a couple hits, put it out. And then it got to a point where I was like ‘All right, I don’t want that.’ So, I was always keeping myself busy” (Age range 21-25, tobacco use during pregnancy). Yet others opted for immediate cessation without a tapering period: “I completely stopped. I don’t know how I did it. But I completely stopped. I bought a pack of cigarettes and went through every single one of them, you know within a couple days. And I just never picked one up after that. I don’t know why. I don’t know how I quit but I just quit” (Age range 21-25, tobacco use during pregnancy).

In addition to reliance on their social circles for support, one participant also mentioned seeking expertise from a health coach for tobacco cessation strategies. However, there was minimal reference of using nicotine replacement products or other medical treatments, and those who tried replacement described it as less successful than self-directed tapering. In this way, participants did not find their obstetric healthcare providers to be an important source of support for behavior change. Although providers offered resources and encouragement, conversations were brief and did not include discussions of alternative approaches other than reduction or abstinence of tobacco use: “I did let my doctor know ‘cause she did. She was like, “Oh, well, if you need a patch or anything — if you need some type of guidance, then we can help you” (Age range 21-25, tobacco use during pregnancy).

#### Postpartum relapse was attributed to difficulty of dual transition into adulthood and parenthood

For participants who endorsed reducing or stopping tobacco use during pregnancy, the majority reported minimal return to prior tobacco use patterns in the postpartum period. They described finding tobacco use more aversive postpartum, with worse cough, stomach pain, headache, and finding the smell and taste less tolerable: “since I had her, I don’t know, my body changed a whole lot. It would cause my chest to hurt. It would ‘cause stomach to hurt, head to hurt. I just stopped that completely” (Age range 16-20, quit nicotine vape upon pregnancy recognition). Interestingly, some individuals describe ongoing craving despite the negative experiences when actually using tobacco postpartum: “I crave a cigarette here and there, but it’s so nasty now, I don’t even want to smoke it, but my body craves it, but my mind doesn’t want it” (Age range 21-25, quit tobacco upon pregnancy recognition). Despite this, many individuals endorse strong motivation to continue maintaining reduced or no tobacco use, citing health concerns for themselves and their offspring as the main reason: “All that being said, with this third and last pregnancy, I feel like I shouldn’t smoke as much, and I shouldn’t go immediately and just go do that. I should try other things, occupy myself other ways before I do that. Sometimes, when it do get too much, I opt out to smoking not a whole one, but at least a half” (Age range 21-25, quit tobacco upon pregnancy recognition).

In contrast, for the minority of participants who reported a return of tobacco use in the postpartum period to prior levels, they described the stress of the postpartum period – especially experiencing it while simultaneously undergoing transition to adulthood – as the main trigger: “I didn’t smoke or nothin’ when I was pregnant, and I was, like, tryin’ to keep it like that, but after I had the baby, I was goin’ through, like, postpartum [blues], and I just thought, uh, ‘Well, I—I’m a new mom. The dad was in jail. And I’m just tryin’ to figure out what I’m gonna do.’ I just graduated from high school” (Age range 16-20, tobacco use during pregnancy). For these individuals, tobacco use provided coping for adulthood and parenthood stress: “At first, it was like I didn’t – I didn’t like – I didn’t like it anymore. It was just like, no, I can’t – this taste is just too weird. I can’t. And then, like, as I, like, inched into it, then I got used to it again, and then I was just like, oh, [audio cut out] I can’t. I just couldn’t just – I just couldn’t no more. I was stressed… It was just like the – like the constant having to get up, being tired all the time. It was just stressful. It was like a whole new transition in my life… It was like a more calm – I would say more calming. Like if he would go to sleep, um, I would just step out of the room and go sit on the porch. It was cold in December. It was freezing. Um, [distorted audio] just sit and just smoke and be like, okay, I have peace and quiet, a little bit of me time again” (Age range 16-20, quit tobacco upon pregnancy recognition). These participants describe the need to cope to maintain functioning as they become primary caregivers to their offspring: “It makes me feel better at the end of the day instead of crying or breaking down or having a mental whatever the heck. Instead of me doing that, it takes my mind off of that, and it makes me focus on something different, completely… Whenever I do smoke in one spot, I focus on that one thing. I’m relaxed enough to handle the next thing instead of being everywhere, crazy, mumbo-jumbo. I can’t think this, I can’t do this, I’m too stressed, too much, it’s too much, it’s too much” (Age range 21-25, quit tobacco upon pregnancy recognition).

## Discussion

This qualitative study of pregnant and postpartum adolescents and young adults identified five themes that describe beliefs, attitudes and experiences related to prenatal tobacco use. Participants recognized tobacco as harmful, perceived it as more harmful than cannabis, and described social networks as catalysts for both initiation and cessation. These themes align with prior research in old pregnant populations. Nonetheless, other findings highlight distinct vulnerabilities in younger pregnant patients. Participants favored self-directed over clinician-supported cessation and attributed postpartum relapse to the compounded stressors of transitioning simultaneously into adulthood and parenthood. our These novel results provide unique insights into needs and preferences for cessation support among younger patients.

### Knowledge of harm did not translate into engagement with cessation resources

Consistent with the broad public awareness of tobacco’s perinatal risks, our participants uniformly viewed tobacco as dangerous for themselves and their offspring^2^. Notably, this knowledge was acquired passively as “common knowledge” rather than through information seeking, and this awareness coexisted with a preference for quitting through personal strategies, such as cutting down, switching to vaping, staying occupied, etc. rather than clinician-offered aids. This coexistence is clinically important because behavioral counseling significantly increases cessation in pregnancy, while health education without counseling has not been proven to be effective^10^. Although our participants received brief provider messages, these communications may not have helped them build the self-efficacy and coping skills that predict long-term abstinence^5^.

### The perception that tobacco is more harmful than cannabis mirrors societal misconceptions

Participants generally ranked tobacco as more dangerous than cannabis and viewed co-use as worse than cannabis alone. This finding is consistent with national data demonstrating that cannabis is perceived as safer than tobacco despite official guidance from resources such as the American College of Obstetricians and Gynecologists, who recommend that pregnant individuals abstain from cannabis due to risks of low birth weight, NICU admission and adverse neurodevelopmental outcomes^18; 19^. Among young adults aged 19-22 years, cannabis use may be as high as 43% and rising social acceptability may deepen the perception of relative safety. There is also evidence that co-use of tobacco and cannabis before and during pregnancy may contribute to continued use and relapse^20^. These gaps between evidence and practice underscore the need for clinicians to counsel on both substances during pregnancy as opposed to tobacco alone.

### Social network influences can be both protective and risk factors

Participants described peers, family, and partners as contributors to initiation but also social support for cessation. The bidirectional role of social networks is well-established in that peer and close-family smoking are some of the most consistent predictors of youth smoking initiation. Social norms have been shown to influence both initiation and cessation^21; 22^. Presence of partners and household members who use nicotine and tobacco is relevant, especially in the postpartum course as living with a smoker is a strong predictor of relapse^2; 5; 23^. Interventions for this population may benefit from engaging partners and family as part of the cessation ecosystem.

### Postpartum relapse highlights unique stressors in young pregnant individuals

Although many participants reported minimal return to smoking, those who relapsed attributed it to stress, fatigue, relationship strain, and needing “me-time” amid a dual transition into adult- and parenthood that adult mothers do not concurrently endure. This qualitative finding is consistent with quantitative evidence that younger maternal age independently predicts relapse. Recent Pregnancy Risk Assessment Monitoring System (PRAMS) data demonstrated that women younger than 20 years old had 1.7-fold higher odds of postpartum relapse and 40% of individuals who quit during pregnancy relapse within a year overall^6^. Established relapse predictors, such as young age, non-breastfeeding, household smoking, and higher psychosocial stressors, map closely onto the stressors reported in this study^5; 23^. Depressive symptoms and stress have been directly linked to relapse in postpartum trials, highlighting the need to address mood and stress in younger age groups^24^.

The strengths of this study include the developmentally focused sample of adolescents and young adults, who comprise an overall understudied group, and inclusion of many participants who identify as Black or African American. In contrast to prior qualitative work on prenatal tobacco use among adolescents, participants were interviewed prospectively, limiting recall bias. Additionally, longitudinal interviews across the perinatal period allowed us to examine if there were changes in beliefs and motivations across pregnancy and in the postpartum. Limitations of the study include recruitment from a single metropolitan health system, lack of participants who identified as Latina or Hispanic, and exclusion of non-English speaking individuals, which limits generalizability.

### Clinical Significance

Our findings suggest that young pregnant patients are receptive to cessation but may not spontaneously engage clinical services and prefer to initially self-direct cessation. Because psychosocial interventions may increase early postpartum abstinence and financial incentives show particular promise, connecting young patients to tailored behavioral support with financial incentives may improve outcomes when supplemented with the current advising at health appointments^11^.

Pharmacotherapy for tobacco use disorder are not recommended by the United States Preventative Task Force^10^ as currently, nicotine replacement therapy in pregnancy is inconclusive although it has shown to be effective in adolescents with tobacco use disorder. Currently, Given the elevated relapse risk in this cohort, relapse-prevention should be discussed later in pregnancy and extend into the postpartum period, with particular focus on household smoking, breastfeeding support, screening for stressors and depressive symptoms with follow-up to provide additional support and treatment^2^.

In conclusion, young pregnant patients possess strong awareness of tobacco and its associated risks, and they may reduce or quit consumption during pregnancy. However, they generally perceive cannabis as a safer option, do not actively engage with clinical resources, and are heavily influenced by social networks and distinct age-relevant environmental stressors. Improving outreach and access to developmentally appropriate cessation and relapse-prevention services in conjunction with optimizing mental health and social environment may improve maternal and infant health in this vulnerable population.

## Data Availability

All data produced in the present study are available upon reasonable request to the authors.

## Notes

### Competing Interest Statement

The authors have declared no competing interest.

### Author Declarations

The University of Pittsburgh Institutional Review Board (IRB) approved all study procedures.

### Funding

- NIDA: R01DA046401

